# N-acetylcysteine as a treatment for substance use cravings: A meta-analysis

**DOI:** 10.1101/2024.05.13.24306839

**Authors:** Emma L. Winterlind, Samantha G. Malone, Michael R. Setzer, Mikela A. Murphy, David Saunders, Joshua C. Gray

**Author notes:** **Corresponding author:** Joshua C. Gray, PhD 4301 Jones Bridge Rd Bethesda, MD 20814. **email addresses:**. **Data availability statement:** The R code used to conduct analyses and create forest and funnel plots can be accessed using this link: https://github.com/ewinterli/NAC-meta-analysis. **Disclosure Statement:** The authors have no commercial or financial conflicts of interests to disclose. **Disclaimer:** The opinions and assertions herein are those of the authors and do not necessarily reflect the official views of the Henry M. Jackson Foundation for the Advancement of Military Medicine, Inc. Moreover, the opinions and assertions herein do not necessarily reflect the official views of the Department of Defense, Uniformed Services University, the National Institute on Alcohol Abuse and Alcoholism, the US Government, and do not imply endorsement by the Federal Government.

## Abstract

N-acetylcysteine (NAC) may serve as a novel pharmacotherapy for substance use and substance craving in individuals with substance use disorders (SUDs), possibly through its potential to regulate glutamate. Though prior meta-analyses generally support NAC’s efficacy in reducing symptoms of craving, individual trials have found mixed results. The aims of the this updated meta-analysis were to (1) examine the efficacy of NAC in treating symptoms of craving in individuals with a SUD and (2) explore subgroup differences, risk of bias, and publication bias across trials. Database searches of PubMed, Cochrane Library, and ClinicalTrials.gov were conducted in June and July of 2023 to identify relevant randomized control trials (RCTs). The meta-analysis consisted of 9 trials which analyzed data from a total of 623 participants. The most targeted substance in the clinical trials was alcohol (3/9; 33.3%), followed by tobacco (2/9; 22.2%) and multiple substances (2/9; 22.2%). Meta-analysis, subgroup analyses, and leave-one-out analyses were conducted to examine treatment effect on craving symptoms and adverse events (AEs). Risk of bias assessments, Egger’s tests, and funnel plot tests were conducted to examine risk of bias and publication bias. NAC did not significantly outperform placebo in reducing symptoms of craving in the meta-analysis (SMD = 0.189, 95% CI = -0.015 - 0.393). Heterogeneity was very high in the meta-analysis (99.26%), indicating that findings may have been influenced by clinical or methodological differences in the study protocols. Additionally, results indicate that there may be publication bias present. Overall, our findings are contrary to those of prior meta-analyses, suggesting limited impact of NAC on substance craving. However, the high heterogeneity and presence of publication bias identified warrants cautious interpretation of the meta-analytic outcomes.

## Introduction

Substance use disorders (SUDs) are one of the most prevalent forms of psychopathology with significant public health disease burden. In 2022 alone, approximately 48.7 million Americans met criteria for any SUD (Substance Abuse and Mental Health Services Administration, 2022). Globally, SUDs contribute to multiple measures of public health disease burden, including years of life lost (YLLs), disability-adjusted life years (DALYs), and years of life lived with disability (YLDs). One global study found that approximately 31.8 million DALYs, 16.7 million YPLs, and 15 million YLDs were attributable to drug use in 2016 (Degenhardt et al., 2018). Additionally, the national economic impact of alcohol and illicit drug misuse is estimated to be $249 and $193 billion, respectively (U.S Department of Health and Human Services, 2023). Despite SUDs posing great public health and economic burdens, current treatments show limited efficacy with relapse rates being as high as 40-60% (Brecht & Herbeck, 2014; McLellan et al., 2000). One particularly strong predictor of relapse is craving, defined as a “strong desire or urge to use” a substance (American Psychiatric Association, 2013, p. 491).

Indeed, a systematic review and meta-analysis by Vafaie & Kober (2022) found that a 1-point increase in drug-related cue and craving indicators more than doubled the odds of substance use or relapse at a later timepoint. Overall, reduction in drug cravings has been proposed as a potentially effective mechanism for reducing substance use (Sayette, 2016; Stone et al., 2024; Witkiewitz et al., 2022).

Individuals with SUD, when presented with drug cues, generally show altered neurobiological craving and reactivity responses in areas of the brain associated with motivational processes, attentional bias, reward-seeking and -processing, and decision-making (Devoto et al., 2020). Importantly, greater activation in these brain regions is associated with poorer treatment outcomes and relapse (MacNiven et al., 2018; Regier et al., 2021). Some pharmacological treatments have been found to reduce both neural craving responses and self-reported craving, thus demonstrating the potential of pharmacological interventions for craving reduction (Cabrera et al., 2016).

N-acetylcysteine (NAC) is an over-the-counter (OTC) antioxidant, approved for the treatment of acetaminophen overdose, which may also serve as an effective pharmacological treatment for craving reduction, primarily due to its effect on glutamate functioning and signaling. The dysregulation of glutamate transmission is one of the most highly researched cellular neuroadaptations associated with chronic substance use (Kalivas, 2009; Volkow et al., 2019). Preclinical models indicate that NAC may reduce drug-seeking behavior by modulating the expression of the glutamate transporter 1 and cysteine-glutamate antiporter, thereby regulating the release of synaptic glutamate in areas of the brain such as the nucleus accumbens core and prefrontal cortex (Baker et al., 2003; Kalivas, 2009; Moro et al., 2020; Roberts-Wolfe & Kalivas, 2015; Zhou & Kalivas, 2008), leading to reduced reinstatement of drug-seeking behavior. These effects have been shown to be long-lasting (Moro et al., 2020; Zhou & Kalivas, 2008).

While several randomized control trials (RCTs) have examined the efficacy of NAC in SUD treatment, including in mitigating craving, many show mixed results (Greenberg et al., 2022). Despite the mixed results found by individual RCTs, meta-analyses support NAC’s efficacy as a pharmacological treatment for craving reduction (Chang et al., 2021; Duailibi et al., 2017). Given the rise in trials focused on the matter, we sought to conduct an updated meta-analysis with the primary aims of (1) examining the efficacy of NAC in treating symptoms of craving in individuals with a SUD and (2) exploring subgroup differences, risk of bias, and publication bias across trials.

## Methods

### Searching Strategy and Study Selection

One author conducted database and registry searches of PubMed, Cochrane Libraries, and ClinicalTrials.gov (https://clinicaltrials.gov). Database searches were conducted on June 20^th^ and July 17^th^, 2023. The following search terms were used: “addiction AND N-acetylcysteine,” “abstinence AND N-acetylcysteine,” “cessation AND N-acetylcysteine,” “craving AND N-acetylcysteine,” “dependence AND N-acetylcysteine,” “substance use disorder AND N-acetylcysteine,” “withdrawal AND N-acetylcysteine,” and “addictive disorder AND N-acetylcysteine.” The search terms were drawn from those of Chang et al. (2021), who, to our knowledge, conducted the most recent meta-analysis on NAC’s efficacy in treating symptoms related to SUDs. The “randomized control trial” and “clinical trial” options in PubMed were used to only retrieve randomized control trials (RCTs). The “search word variations” option in Cochrane Libraries was used to broaden the search scope. No language restrictions were applied. Two authors independently reviewed database and registry search results for the study selection process and a third author served as a third reviewer in the case of unresolvable discrepancies between the primary reviewers. Trial inclusion judgments were based on the reviewers’ individual assessments of whether trials fit the inclusion and exclusion criteria. Initial agreement between the primary reviewers at the full-text review stage was 87.1% (Cohen’s Kappa [κ] = 0.746), indicating substantial agreement (McHugh, 2012). After discussion, 100% agreement was reached (κ = 1.0). All authors reviewed and agreed upon the final trials included in the meta-analysis.

### Inclusion and Exclusion Criteria

We used the following inclusion criteria: (a) randomized, placebo-controlled trials with no crossover; (b) standardized assessment(s) of craving using validated measures; (c) only adjuvant treatments to NAC allowed were psychosocial or behavioral therapies (e.g., cognitive behavioral therapy, contingency management); and (d) include only participants who meet DSM-IV, DSM-5, or DSM-5-TR criteria for any SUD. An exception to criterion D was made for tobacco use trials; to be included in our meta-analysis, trials examining the efficacy of NAC as a treatment for tobacco use had to include an eligibility criterion specifying that participants had to smoke ≥10 cigarettes per day. We excluded pre-clinical trials, case reports, trial protocols, systematic reviews or meta-analyses, non-controlled trials, trials with adjuvant pharmacological or non-psychosocial or -behavioral treatments to NAC, and trials not assessing substance use and craving. We only allowed for psychosocial or behavioral therapies as adjuvant treatments to NAC because these are the standard, evidence-based treatments for SUD (National Institute on Drug Abuse, 2011) and, in theory, would likely be the ones administered alongside NAC in clinical practice.

### Data Extraction

The primary outcome was craving. Baseline and endpoint craving data were independently extracted and then compared by two authors to ensure accuracy. If a trial reported scores at more than one time-point after baseline we used the scores corresponding to the longest time period for which participants were blinded.

If potentially eligible trials did not include sufficient data or information for inclusion in our meta-analysis, the authors were contacted three times to retrieve the data or information. Insufficient data or information for inclusion in the meta-analysis include not reporting post-intervention scores (means and standardized deviation [SD] or standard error [SE]) for the craving measure(s) used in their trial or not posting trial results to ClinicalTrials.gov (if the trial was only posted to ClinicalTrials.gov). If the authors did not respond after three contact attempts (k = 3) or were unable to provide the data requested (k = 2), the study was excluded (k = 5). One study (Roten et al., 2013) did not include follow-up data in their publication but provided it upon request. Trial characteristics such as author names, inclusion criteria, demographics, substance treated, measure(s) used, and intervention details were independently extracted by two authors and then compared to ensure accuracy. Trial characteristics are presented in Table 1. The total number of trials included in the present meta-analysis (k = 9) was slightly lower than the generally recommended number (k = 10; Borenstein & Higgins, 2013; Richardson et al., 2019).

**Table 1.**
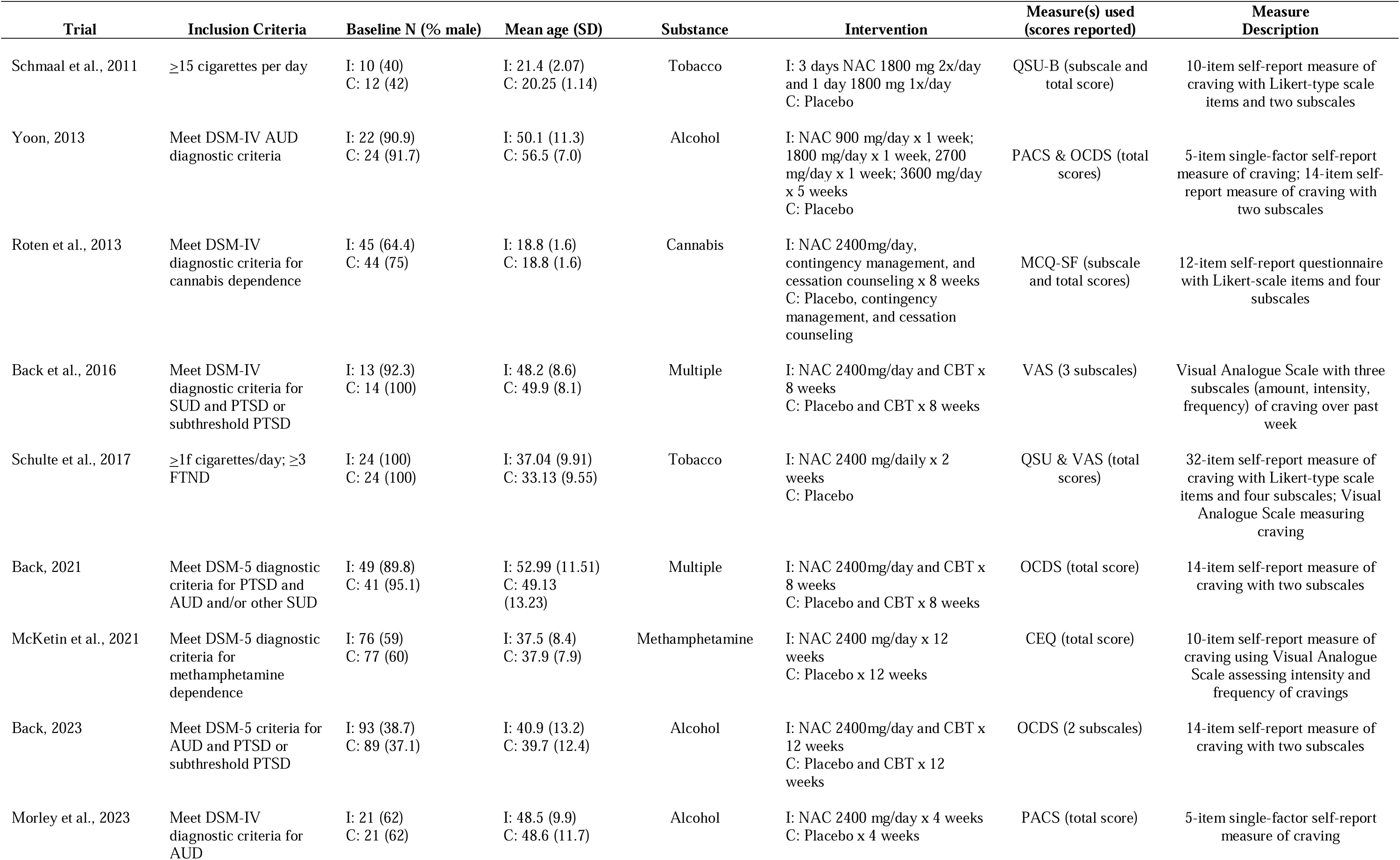
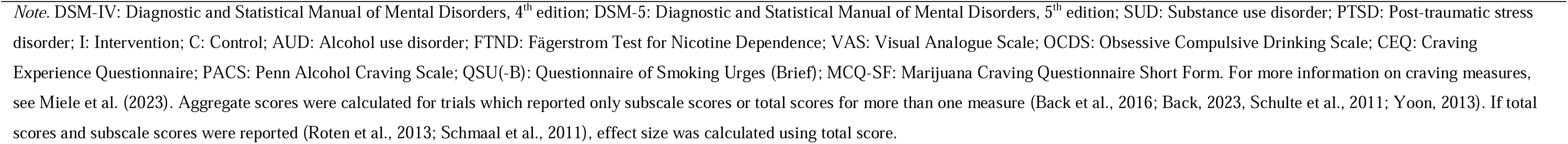
Trial Characteristics.

| **Trial** | **Inclusion Criteria** | **Baseline N (% male)** | **Mean age (SD)** | **Substance** | **Intervention** | **Measure(s) used (scores reported)** | **Measure Description** |
| --- | --- | --- | --- | --- | --- | --- | --- |
| Schmaal et al., 2011 | ≥15 cigarettes per day | I: 10 (40)<br>C: 12 (42) | I: 21.4 (2.07)<br>C: 20.25 (1.14) | Tobacco | I: 3 days NAC 1800 mg 2x/day and 1 day 1800 mg 1x/day<br>C: Placebo | QSU-B (subscale and total score) | 10-item self-report measure of craving with Likert-type scale items and two subscales |
| Yoon, 2013 | Meet DSM-IV AUD diagnostic criteria | I: 22 (90.9)<br>C: 24 (91.7) | I: 50.1 (11.3)<br>C: 56.5 (7.0) | Alcohol | I: NAC 900 mg/day x 1 week; 1800 mg/day x 1 week; 2700 mg/day x 1 week; 3600 mg/day x 5 weeks<br>C: Placebo | PACS & OCDS (total scores) | 5-item single-factor self-report measure of craving; 14-item self-report measure of craving with two subscales |
| Roten et al., 2013 | Meet DSM-IV diagnostic criteria for cannabis dependence | I: 45 (64.4)<br>C: 44 (75) | I: 18.8 (1.6)<br>C: 18.8 (1.6) | Cannabis | I: NAC 2400mg/day, contingency management, and cessation counseling x 8 weeks<br>C: Placebo, contingency management, and cessation counseling | MCQ-SF (subscale and total scores) | 12-item self-report questionnaire with Likert-scale items and four subscales |
| Back et al., 2016 | Meet DSM-IV diagnostic criteria for SUD and PTSD or subthreshold PTSD | I: 13 (92.3)<br>C: 14 (100) | I: 48.2 (8.6)<br>C: 49.9 (8.1) | Multiple | I: NAC 2400mg/day and CBT x 8 weeks<br>C: Placebo and CBT x 8 weeks | VAS (3 subscales) | Visual Analogue Scale with three subscales (amount, intensity, frequency) of craving over past week |
| Schulte et al., 2017 | ≥1f cigarettes/day; ≥3 FTND | I: 24 (100)<br>C: 24 (100) | I: 37.04 (9.91)<br>C: 33.13 (9.55) | Tobacco | I: NAC 2400 mg/daily x 2 weeks<br>C: Placebo | QSU & VAS (total scores) | 32-item self-report measure of craving with Likert-type scale items and four subscales; Visual Analogue Scale measuring craving |
| Back, 2021 | Meet DSM-5 diagnostic criteria for PTSD and AUD and/or other SUD | I: 49 (89.8)<br>C: 41 (95.1) | I: 52.99 (11.51)<br>C: 49.13 (13.23) | Multiple | I: NAC 2400mg/day and CBT x 8 weeks<br>C: Placebo and CBT x 8 weeks | OCDS (total score) | 14-item self-report measure of craving with two subscales |
| McKetin et al., 2021 | Meet DSM-5 diagnostic criteria for methamphetamine dependence | I: 76 (59)<br>C: 77 (60) | I: 37.5 (8.4)<br>C: 37.9 (7.9) | Methamphetamine | I: NAC 2400 mg/day x 12 weeks<br>C: Placebo x 12 weeks | CEQ (total score) | 10-item self-report measure of craving using Visual Analogue Scale assessing intensity and frequency of cravings |
| Back, 2023 | Meet DSM-5 criteria for AUD and PTSD or subthreshold PTSD | I: 93 (38.7)<br>C: 89 (37.1) | I: 40.9 (13.2)<br>C: 39.7 (12.4) | Alcohol | I: NAC 2400mg/day and CBT x 12 weeks<br>C: Placebo and CBT x 12 weeks | OCDS (2 subscales) | 14-item self-report measure of craving with two subscales |
| Morley et al., 2023 | Meet DSM-IV diagnostic criteria for AUD | I: 21 (62)<br>C: 21 (62) | I: 48.5 (9.9)<br>C: 48.6 (11.7) | Alcohol | I: NAC 2400 mg/day x 4 weeks<br>C: Placebo x 4 weeks | PACS (total score) | 5-item single-factor self-report measure of craving |

### Risk of Bias Assessments

Two authors independently assessed the methodological quality of each study based on the revised Cochrane Risk of Bias (RoB) tool for randomized control trials (Higgins et al., 2011) before coming together to reach consensuses. Another author was assigned as a third reviewer in the case of unresolvable discrepancies between the primary reviewers; however, all discrepancies were able to be resolved through discussion between the two primary reviewers.

### Statistical Analysis

The meta-analysis was conducted in accordance with PRISMA guidelines (Page et al., 2021). The meta-analysis was not pre-registered and no study protocol was prepared for publication. The meta-analysis and subgroup analyses were all performed using the metafor package (Viechtbauer, 2010) in R version 4.3.3.

Cohen’s *d*, a type of standardized mean difference (SMD), was independently calculated for each study by two authors and then compared to ensure accuracy. Several trials reported total scores for more than one measure or only subscale scores, but not total score, for a measure (Back et al., 2016; Back, 2023; Schulte et al., 2017; Yoon, 2013). Rather than selecting only one measure or subscale to include in our meta-analysis, or treating each measure as an independent study, we calculated an aggregated score using two methods: by aggregating effect sizes in the MAd package in R (Del Re & Hoyt, 2014) and by manually totaling scores and dividing by the number of measures or subscales. Both methods resulted in the same aggregated effect size for all the relevant trials. Logarithmic risk ratios (logRRs) were calculated to compare reports of adverse events (AEs) between placebo and intervention groups. Confidence intervals (CIs) were automatically calculated for SMDs and RRs by the metafor package when trials were fitted to a model. The package also calculates several heterogeneity statistics; we focus on the *I^2^* statistic when reporting the results. The random effects (RE) model was chosen to best account for between-study variability and heterogeneity.

In the case of one study (McKetin et al., 2021), Cohen’s *d* for the craving measure was found to be null and, thus, the standard error (SE) was also found to be null. To account for this, we transformed the SE to a small decimal (0.001) while maintaining the null effect size. To account for any bias this may introduce, we conducted a subgroup analysis which excluded this trial but maintained all other trials included in the meta-analysis (SA1). Moreover, we were interested to see whether the substance type treated may impact the effect of treatment. To examine this, we conducted a subgroup analysis (SA2) similar to the one conducted by Chang et al. (2021) which focused on trials examining alcohol to assess whether NAC had a greater treatment effect for alcohol, the substance type with the most recent and greatest number of trials (Back, 2023; Morley et al., 2023; Yoon, 2013), than for other substances. Finally, we conducted a subgroup analysis (SA3) examining group differences in logRRs for AEs. We report on all meta-analytic and subgroup findings.

First, the meta-analysis including all nine trials was conducted, and alongside it, the first subgroup analysis excluding McKetin et al., 2021 (SA1; Figure 2). The second subgroup analysis examining only trials treating alcohol use was then conducted (SA2; Figure 3). Finally, the subgroup analysis of risk ratios was conducted to compare risk of AEs between placebo and intervention groups (SA3; Figure 4). Leave-one-out-analyses (LOOAs) were conducted to examine the influence of each study on the overall outcomes of the meta-analysis and the subgroup excluding McKetin et al. (2021). Additionally, publication bias was assessed through the use of Begg’s funnel plots and Egger’s tests (Egger et al., 1997).

## Results

### Trial Characteristics

Figure 1 displays the literature screening and selection process. The initial search yielded 955 records, 636 of which were excluded for being duplicates, leaving a total of 319 records for title and abstract screening. Another potentially eligible study (Morley et al., 2023) was identified by one of the authors after the initial database searches were completed; the study was added to the review process, thus leaving a total of 320 records for title and abstract screening.

**Figure 1.**
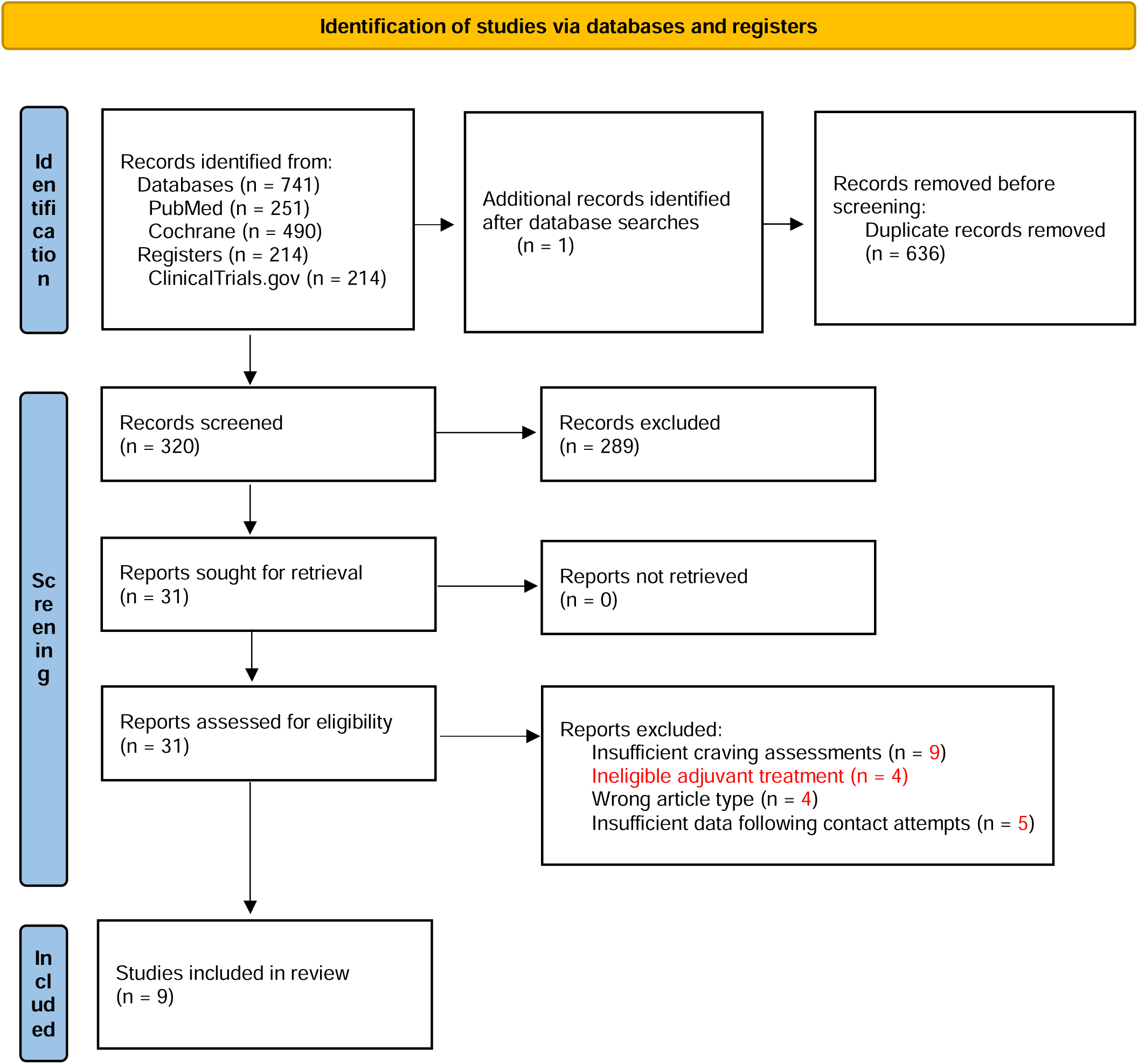
PRISMA flow diagram depicting study selection process

After titles and abstracts were screened for relevance, a total of 31 records remained for full-text review. Two trials were excluded for including a pharmacological adjunct to NAC and two for including another ineligible adjuvant treatment, four for being the wrong article type, nine for insufficient assessment(s) of craving (e.g., use of unstandardized assessments, no assessments, or assessment only at baseline), and five for having insufficient data following the contact attempts. The two trials excluded for including other non-pharmacological ineligible adjuvant treatments were associated trials with the same dataset. A total of nine trials were included in the final meta-analysis. The trials were published between 2013 and 2023 and had sample sizes ranging from 22 to 153, with a total of 699 participants at baseline. However, due to attrition within the trials, the total number of participants analyzed was 623.

The measures of craving used were all self-report. The Craving Experiences Questionnaire (CEQ), Obsessive Compulsive Drinking Scale (OCDS) and Penn Alcohol Craving Scale (PACS) assessed generalized craving (i.e., general senses of craving, usually across the past week). The Visual Analogue Scale (VAS), employed by Back et al. (2016) and Schulte et al. (2017), can be used to measure both generalized or acute (i.e., current or immediate sense of craving) craving. Back et al. (2016) used the VAS in a generalized fashion, assessing intensity, frequency, and amount of craving over the past week, whereas Schulte et al. (2017) did not specifically document in what fashion they used the scale. The Questionnaire of Smoking Urges (QSU) measures acute craving and the Marijuana Craving Questionnaire (MCQ) measures both acute and generalized craving. All measures except the VAS and CEQ are explicitly designed to measure craving for a specific substance. None of the trials used cue-induction prior to craving assessment and all measures were administered to participants in-person during follow-ups. For more information on these and other craving measures, see Miele et al. (2023). Table 1 describes trial characteristics in further detail.

### Craving Symptoms

Baseline data were reported in all trials. Follow-up data were reported in eight trials; one study did not report follow-up data but provided these data upon request (Roten et al., 2013). We first conducted the meta-analysis and SA1. In the meta-analysis, we found that NAC did not significantly outperform placebo in reducing symptoms of craving (SMD = 0.189, 95% CI = - 0.015 - 0.393). Very high heterogeneity was observed across trials (*I*^2^ = 99.26%). The results of SA1 further confirmed the findings of the meta-analysis (SMD = 0.217, 95% CI = -0.012 - 0.446, *I*^2^ = 97.33%). A test for group differences (meta-analysis versus subgroup) identified no significant differences between the groups (*p* = 0.87), indicating little to no impact of including the null effect size study on the overall outcome of the meta-analysis. Figure 2 depicts a forest plot with outcomes of the RE models for the meta-analysis, subgroup analysis, and the test for subgroup differences.

**Figure 2.**
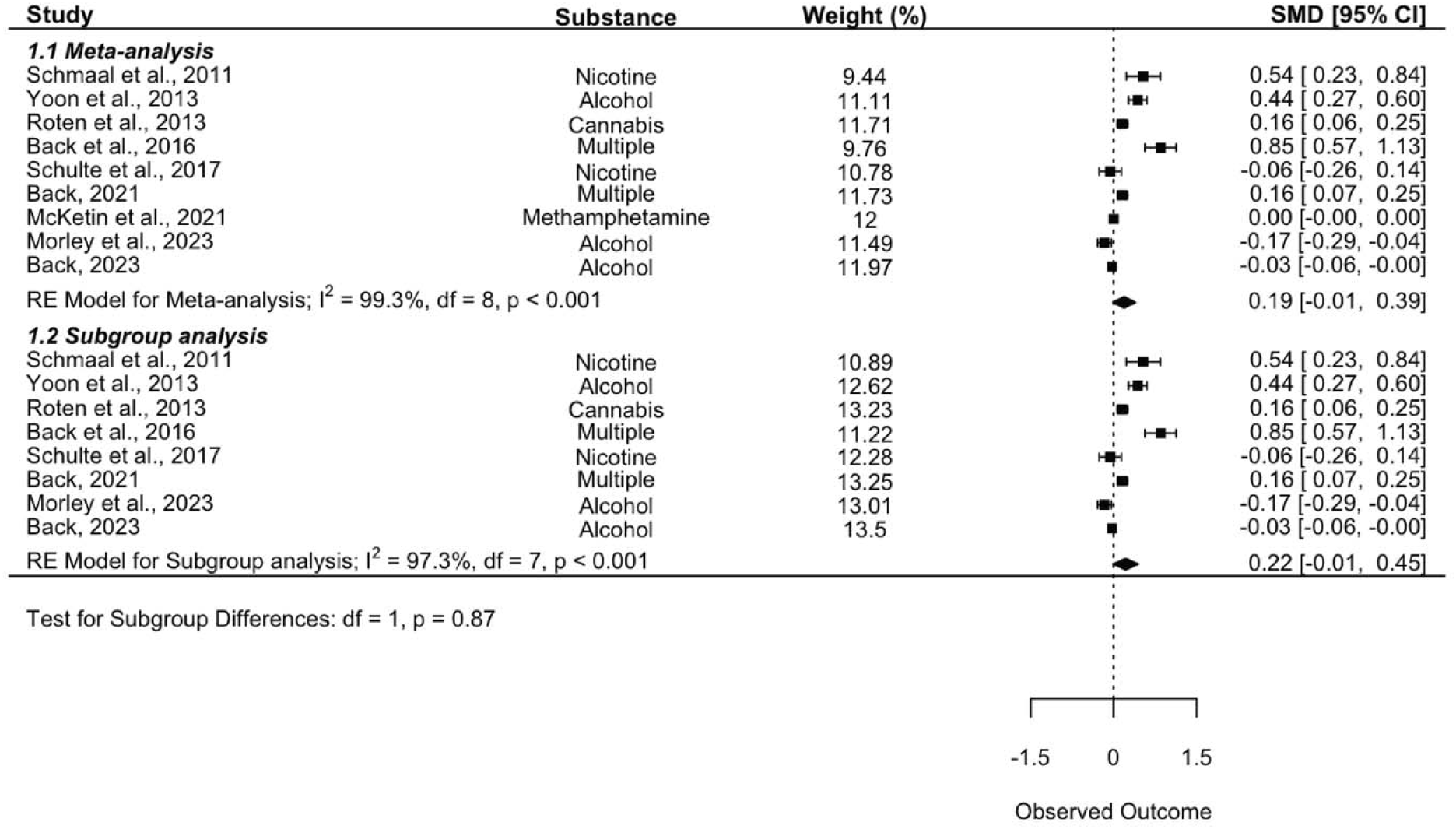
Meta-analysis forest plot. The plot depicts a meta-analysis and subgroup analysis examining the overall effect of NAC on the primary outcome, as well as a test for subgroup differences. Note that values above the line of no effect indicate favorability towards the intervention (NAC) group.

SA2, the subgroup analysis examining only trials which were targeting alcohol use, also yielded insignificant results (SMD = 0.074, 95% CI = -0.277 - 0.425, *I*^2^ = 97.11%). The test for subgroup differences (alcohol trials versus all other trials) identified no significant differences between the groups (*p* = 0.43). Figure 3 depicts a forest plot with outcomes of the RE models for SA2 and test for subgroup differences.

**Figure 3.**
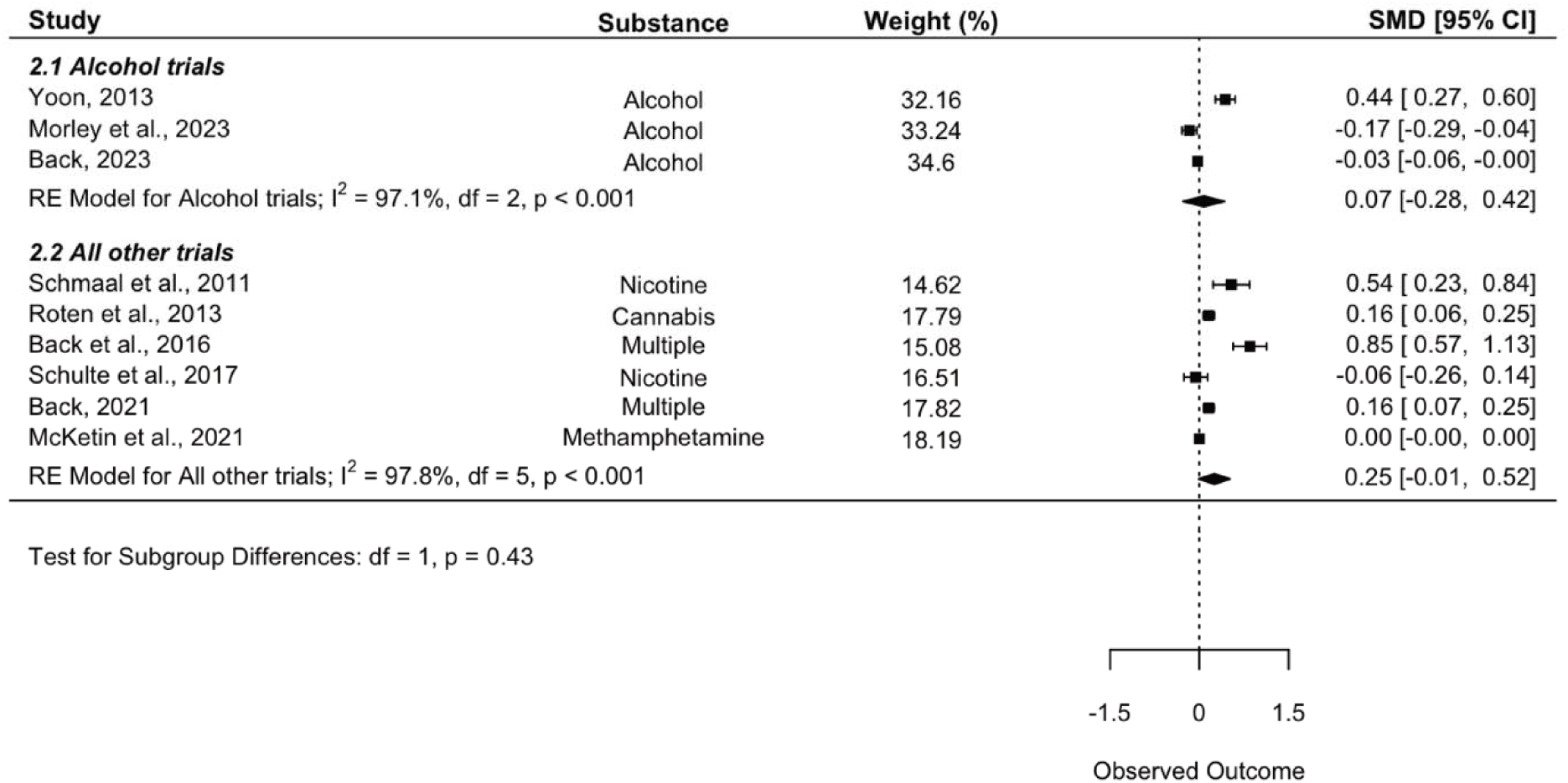
Alcohol trials subgroup forest plot. The plot depicts a subgroup analysis comparing trials examining alcohol to all other trials. Note that values above the line of no effect indicate favorability towards the intervention (NAC) group.

LOOAs were conducted to examine the influence of each study on the outcomes of the meta-analysis and SA1. The LOOAs identified Morley et al. (2023) as a significantly influential study in both the meta-analysis (SMD = 0.235, 95% CI = 0.025 - 0.444, *p* = 0.03) and SA1 (SMD = 0.273, 95% CI = 0.040 - 0.506, *p* = 0.02), indicating that if this study were to be excluded from the meta-analysis or subgroup analysis, it would change the overall results to indicate that NAC did significantly outperform placebo. The LOOA on SA1 also identified Back (2023) (SMD = 0.257, 95% CI = 0.002 - 0.511, *p* = .049) and Schulte et al. (2017) (SMD = 0.257, 95% CI = 0.006 - 0.508, *p* = 0.045) as significantly influential trials.

### Adverse Events

Eight trials reported on AEs experienced by participants, while one did not (Roten et al., 2013). There were 16 serious adverse effects reported in the experimental groups across trials, only one of which was conservatively estimated to have a possible relation to the intervention (syncopal episode; Back et al., 2016). Gastrointestinal upset (e.g., stomach cramps and discomfort, diarrhea, nausea) and headache were the most commonly reported non-severe AEs.

SA3 was conducted to examine group differences in RRs for AEs. Though eight trials reported AEs, one study did not report occurrences of AEs between intervention and control group (Back et al., 2016). Another trial reported a greater number of adverse events than the number of participants for both groups (McKetin et al., 2021) and another reported zero AEs for both groups (Schulte et al., 2017), thereby making the calculation of risk ratios for these trials impossible. Of note, McKetin et al. (2021) appeared to have employed the most thorough assessment of AEs out of all eight trials, coding AEs by Systems Organ Class according to the Medical Dictionary for Regulatory Activities (MedDRA; Jaasu et al., 2018). Only one other study mentioned a standardized assessment of AEs (Back et al., 2016), whereas all other trials either did not report how they collected data on AEs or simply mentioned asking participants if they experienced any side effects. All in all, four trials were excluded from this subgroup analysis (Back et al., 2016; McKetin et al., 2021; Roten et al., 2013; Schulte et al., 2017), leaving a total of five trials to be included. The results indicate no significant differences in risk for AEs by group (NAC versus placebo; logRR = 0.90, 95% CI = 0.68 - 1.19). Figure 4 depicts a forest plot with outcomes of the RE model for the AE subgroup analysis.

**Figure 4.**
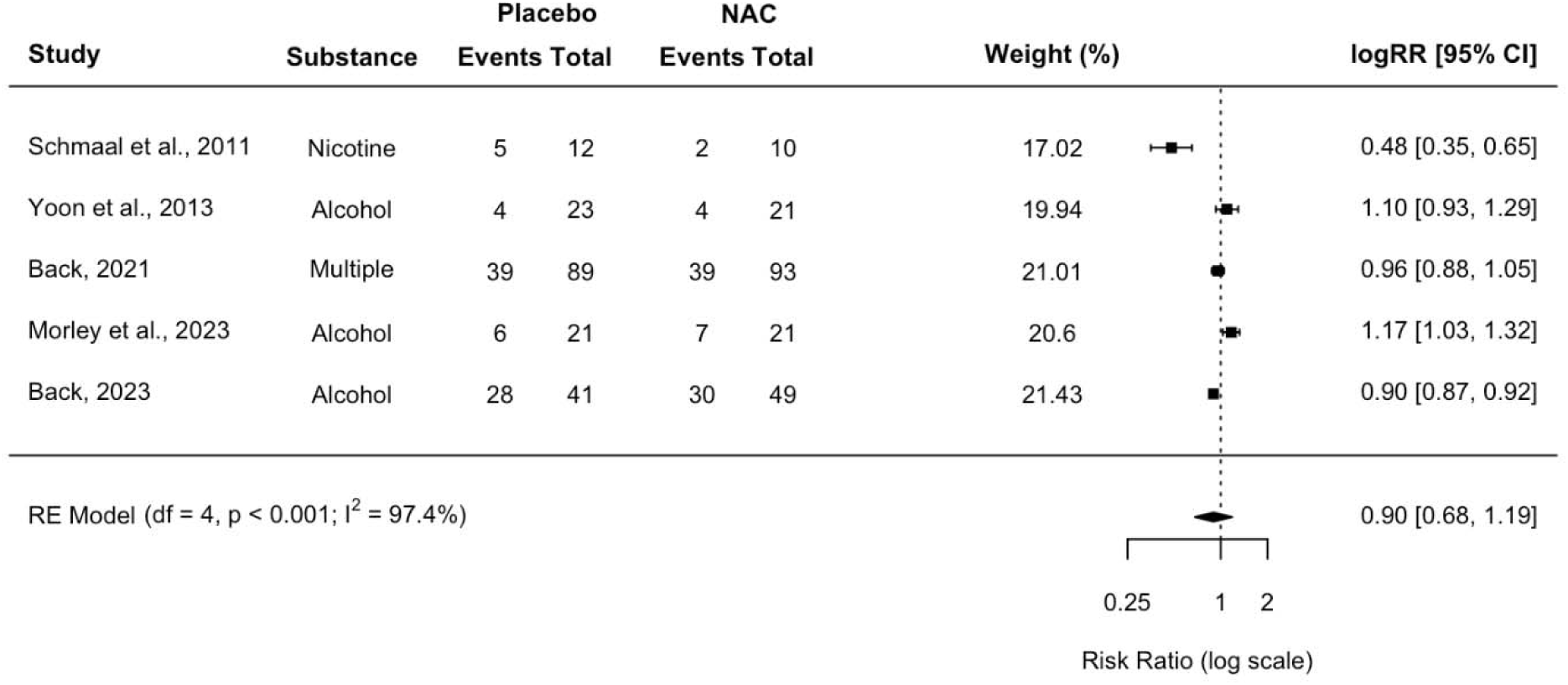
Log risk ratio forest plot. The plot depicts a subgroup analysis of risk of adverse events. Note that values below the line of no effect indicate lower risk of AEs for the intervention (NAC) group.

### Risk of Bias and Publication Bias

The methodological quality (i.e., risk of bias), as assessed by the Cochrane Risk of Bias tool (Higgins et al., 2011), of the included trials are described in Table 2 and Figure 6. Regarding overall bias, seven trials were rated as high risk and two as some concerns. The third domain of the Cochrane RoB tool, missing outcome data, was a consistent source of high bias across trials. Additional sources of bias identified by the reviewers include underpowered trials and lack of pre-specified plans of analysis.

**Table 2.** Risk of Bias Judgments.

| **Trial** | **Domain 1** | **Domain 2** | **Domain 3** | **Domain 4** | **Domain 5** | **Overall judgment** |
| --- | --- | --- | --- | --- | --- | --- |
| Schmaal et al., 2011 | Low | Low | Low | Low | Some concerns | Some concerns |
| Yoon, 2013 | Some concerns | Low | High | Low | Some concerns | High |
| Roten et al., 2013 | Some concerns | Low | High | Some concerns | Some concerns | High |
| Back et al., 2016 | Low | High | High | Low | Low | High |
| Schulte et al., 2017 | Some concerns | High | High | Low | Some concerns | High |
| Back, 2021 | Low | Low | High | Low | Low | High |
| McKetin et al., 2021 | High | Some concerns | Low | Some concerns | Low | High |
| Back, 2023 | Low | Low | High | Low | Low | High |
| Morley et al., 2023 | Low | Low | Low | Low | Some concerns | Some concerns |

*Note.* Domain 1: Bias arising from the randomization process; Domain 2: Bias due to deviations from intended intervention; Domain 3: Bias due to missing outcome data; Domain 4: Bias in measurement of the outcome; Domain 5: Bias in selection of the reported result

Two Begg’s funnel plots were created to assess publication bias of the included trials, one for the meta-analysis and one for SA1. Since the plots looked virtually identical, only the plot which includes all trials is presented (Figure 5). Both funnels exhibited asymmetry, potentially indicating publication bias. Additionally, Egger’s tests (Egger et al., 1997) were conducted on each funnel plot to further assess publication bias. If significant, results of Egger’s tests indicate that publication bias favoring statistically significant or positive trials may be more likely to be present. Both tests indicated statistically significant asymmetry (*p*s < .05), further indicating publication bias. These findings are contrary to those of a prior meta-analysis on the subject (Duailibi et al., 2017).

**Figure 5.**
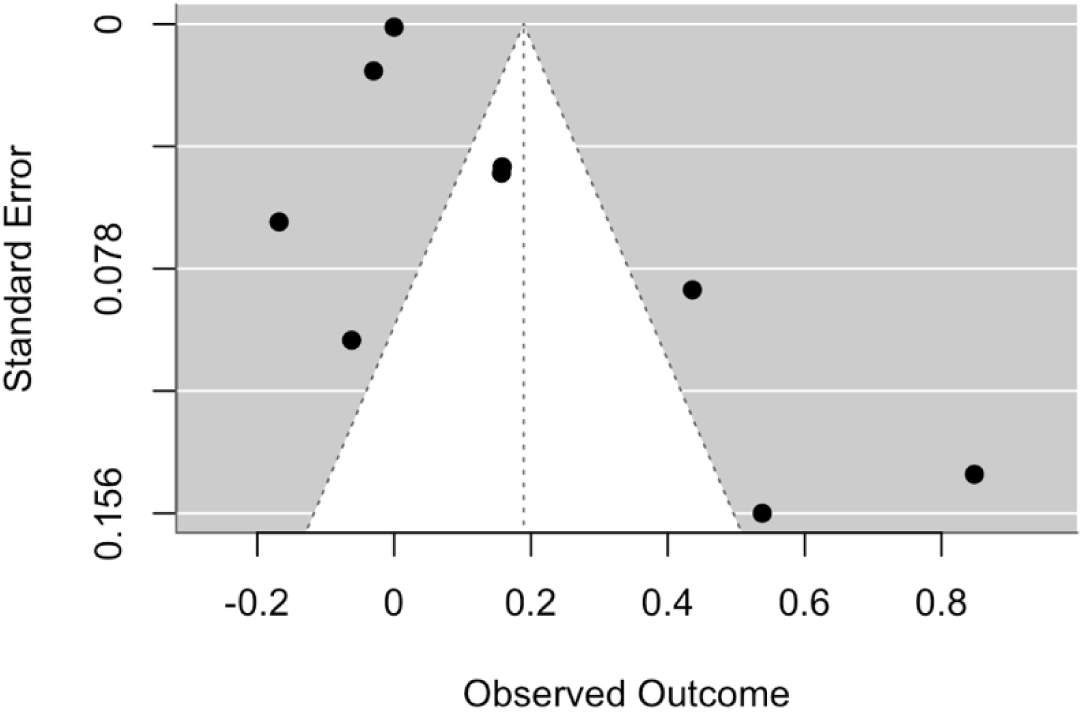
Begg’s funnel plot. The plot depicts the outcome of a funnel plot including all trials in the meta-analysis.

**Figure 6.**
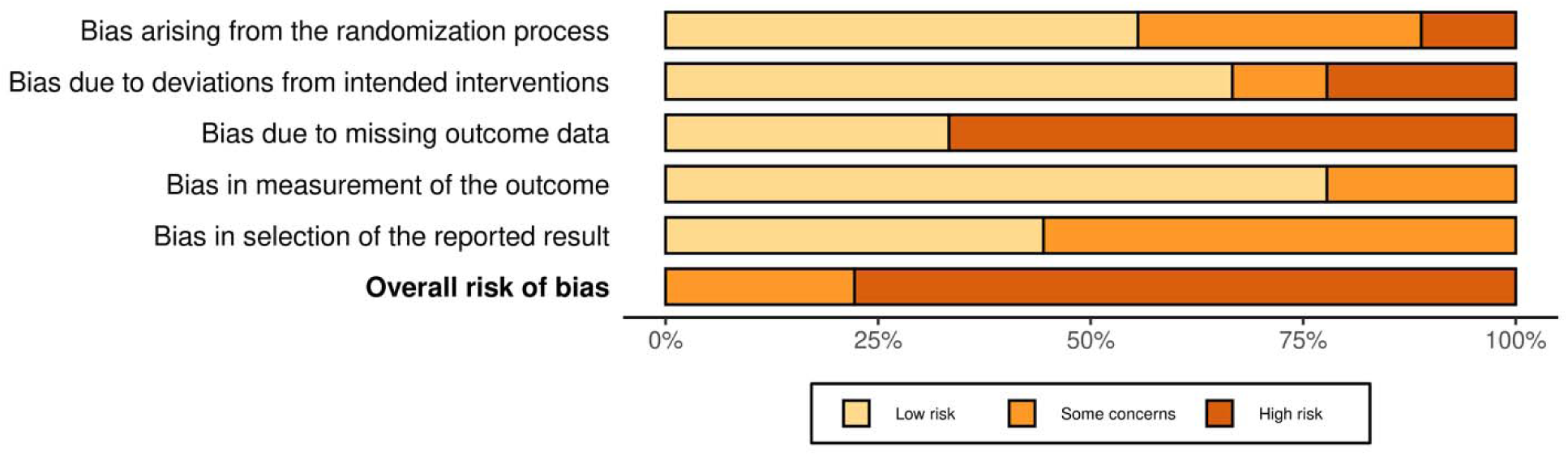
Risk of Bias Bar Plot. The figure depicts overall and domain-specific risk of bias judgments each included study.

## Discussion

Our meta-analysis found that NAC was not superior to placebo in reducing cravings in individuals with SUD. These findings contrast with those of Chang et al. (2021) and Duailibi et al. (2017), who both found that NAC was superior to placebo in treating substance use cravings and other symptoms associated with SUD (e.g., withdrawal and depression). However, our meta-analysis included several more recent and large trials (Back, 2021, 2023; McKetin et al., 2021; Morley et al., 2023), most of which had smaller treatment effect sizes than those included in the prior meta-analyses.

Smaller, earlier trials are more prone to inflate treatment effects (i.e., ‘small-study effects), potentially as a result of lesser methodological quality (Dechartres et al., 2013). This may increase the likelihood of publication bias, the tendency for trials with statistically significant or positive results to be published over trials with null findings (Dalton et al., 2016).

Indeed, the two trials with the largest effect sizes included in this meta-analysis (Back et al., 2016; Schmaal et al., 2011) also had the smallest sample sizes (*N*s = 27 and 22, respectively). We identified significant asymmetry in the funnel plots, a tool considered to be effective in detecting small-study effects (Egger et al., 2003), though it may be considered unreliable when used in meta-analyses with less than 10 trials (Dalton et al., 2016). While it appears likely that publication bias primarily influenced the asymmetry, it is worth noting that publication bias is only one of many potential reasons for funnel plot asymmetry (e.g., low methodological quality, selection biases, true heterogeneity, data irregularities, artifact; Egger et al., 1997).

This meta-analysis is not without its limitations. The most notable limitation was the relatively low number of trials included (k = 9), which is slightly below the recommended number of trials for a meta-analysis (k = 10; Borenstein & Higgins, 2013; Richardson et al., 2019). This was in part due to the number of studies for which we were unable to acquire the sufficient data to include in the meta-analysis (k = 5). Crossover trials were also excluded after considering the documented risks and limitations of including such trial designs in meta-analyses. Such risks and limitations include period and carryover effects, insufficient within-individual treatment comparisons, higher dropout, and heightened impact of missing data on trial outcomes (Li et al., 2015). However, only two small (*N*s = 12 and 23) trials were excluded for the sole reason of being of a crossover study design. The lower number of trials included in the meta-analysis also impacted our ability to explore potential sources of heterogeneity with statistical confidence.

Focusing solely on craving reduction may also have limited our conclusions. We chose to assess craving reduction because of evidence suggesting that it is a notable mechanism of behavior change leading to reductions in substance use (Stone et al., 2024), a finding which has been translated across various substance types, including alcohol, cannabis, and cocaine (Sayette, 2016; Stone et al., 2024; Witkiewitz et al., 2022). Moreover, our meta-analysis specifically sought to expand upon the prior meta-analyses conducted by Chang et al. (2021) and Duailibi et al (2017), both of which which shared a primary outcome of craving reduction. Still, we recognize that craving reduction is only one of several important areas of addiction management and, as such, focusing solely on craving reduction as the primary outcome of the meta-analysis limits our ability to understand how NAC may impact other important areas of SUD management and treatment, such as substance use reduction and functional impairment improvement. Moreover, examining other treatment areas such as substance use reduction may have allowed for greater analytical exploration and insight into substance-specific treatment effects of NAC. To expand upon the findings of the present meta-analysis, we suggest that future meta-analyses explore the efficacy of NAC in treating various areas of SUD management, including quantity and frequency of use, abstinence, and relapse prevention.

The high levels of heterogeneity across trials utilized in the meta-analysis possibly limits our conclusions. The nine trials included in this analysis often did not share methodologies or measurements of craving, making it difficult to draw accurate conclusions from the aggregate results regarding the utility of NAC as a treatment for SUD cravings. Particularly, the variability in craving measures used and the use of aggregated craving scores may have impacted findings. Additionally, because trials vary in the SUD treated, it is difficult to determine whether NAC serves as a more effective treatment for certain substances over others. However, the findings from the subgroup analysis examining only alcohol-focused trials support the findings of Chang et al. (2021), who similarly conducted a subgroup analysis of alcohol-focused trials and found a nonsignificant treatment effect of NAC on craving, and Duailibi et al. (2017), who found that substance type did not moderate treatment effects.

Regarding the high heterogeneity, it is possible that the *I*^2^ estimate for the present meta-analysis was inflated and results were more homogenous than predicted. Evidence suggests that *I*^2^ demonstrates considerable bias when there are few trials included in analyses. When there is minimal true heterogeneity, this measure often increases it artificially, but when there is large true heterogeneity, this measure often decreases it artificially (Von Hippel, 2015). Because of this, the *I*^2^ heterogeneity statistic should be interpreted cautiously and the real extent to which heterogeneity impacted our results is unclear.

Another important consideration which may have impacted results is the quality of the NAC product itself. It is possible that there were variations in quality (e.g., purity, potency, preparation) and regulatory status of the NAC products used in the included trials of this meta-analysis. The quality of NAC products was not systematically assessed in the present study or in prior meta-analyses (Duailibi et al., 2017; Chang et al., 2021). Though NAC has undergone a rigorous regulatory process in order to become a FDA-approved medication for the treatment of acetaminophen overdose, it is also a widely available OTC dietary supplement for which the FDA exercises limited enforcement discretion (U.S. Department of Health and Human Services, 2022). Since OTC supplements are unregulated by the FDA, without guarantee of purity or medication-substance content, the effectiveness of a given supplement may be limited. As such, it is recommended that NAC products are sourced from suppliers which meet United States Pharmacopeia (USP) standards (Tomko et al., 2018).

This meta-analysis corroborated the findings of prior large RCTs with null results, despite prior meta-analytic evidence indicating that NAC is a promising treatment option for substance craving. Large RCTs will enhance generalizability, reduce potential bias, and amplify study power to detect real effects. Similarly, considering homogeneity in trial design (e.g. trial protocols, outcome measures, and the SUD treated) will enhance the ability of RCTs to evaluate the efficacy of NAC as a SUD treatment more accurately. Additionally, researchers should publish negative findings when applicable to help prevent publication bias. These remediations will improve the ability of future meta-analyses to clarify NAC treatment effects and identify moderators of treatment effect.

## Data Availability

The R code used to conduct analyses and create forest and funnel plots can be accessed using this link: https://github.com/ewinterli/NAC-meta-analysis

